# Deep Learning based CT-scan Coronary Artery Segmentation and Calcium Scoring

**DOI:** 10.1101/2024.09.06.24313174

**Authors:** Sai Koundinya Upadhyayula

## Abstract

Coronary artery disease (CAD), primarily driven by atherosclerosis, poses significant health risks, contributing to a rising mortality rate globally. This study introduces a deep learning framework designed for the automated segmentation of coronary arteries and quantification of coronary artery calcium (CAC) from CT scans, facilitating improved risk stratification in patients. Leveraging data from the National Lung Screening Trial, we developed a three-step model that includes heart localization, coronary calcium segmentation, and calcium scoring. Various configurations of the UNet architecture were employed, with the Extended UNet utilizing an autoencoder achieving the highest validation performance, reflected by an Intersection over Union (IoU) score of 0.78 and an F1 score of 0.83.

The model’s efficacy was validated against manually segmented masks, showcasing its potential for accurate risk assessment based on CAC scores. This automated approach significantly reduces the time and expertise required for traditional calcium scoring, enabling rapid and reliable assessments in clinical settings. Our findings indicate that the deep learning system can effectively classify patients into risk categories, underscoring its potential utility in enhancing the management of CAD and improving patient outcomes. This research highlights the feasibility of integrating advanced computational techniques into routine clinical practice, paving the way for more efficient cardiovascular risk stratification.

## 1. Introduction

The CAD is a cardiovascular disorder occurring due to atherosclerosis or atherosclerotic occlusions of the coronary arteries. The disease process is characterized by stable angina, progressing to unstable angina and myocardial infarction (MI)/ sudden cardiac death precipitated by arrhythmogenic ischemic myocardium. The contribution of CAD to total deaths and disease burden in India has almost doubled since 1990. With an estimated 24 million patients suffering from CAD, and an estimated mortality of 2.8 million patients in 2016.

The incipience of the disease is with endothelial dysfunction and altered permeability, which leads to diffusion of low-density lipoproteins through the arterial wall, into the intima. The cholesterol rich LDL is then oxidized, serving as a powerful chemoattractant for leukocytes. Macrophage mediates phagocytosis of these compounds leads to further release of inflammatory mediators and the formation of foam cells. These foamy cells replicate and organize to form lesions called fat streaks. This is the earliest form of lesions visualized in atherosclerosis. The formation of these lesions signals the proliferation of smooth muscle cells, causing the fatty streak to enlarge. The SMCs produce a large amount of extracellular matrix, which forms the scaffolding of the atherosclerotic plaque. Chronic deposition of LDL and inflammatory/fibrotic remodelling of the lesion narrows the arterial lumen and precipitates myocardial ischemia at >60% occlusion. The atherosclerotic plaques, being rich in lipids, cholesterol esters and necrotic cell debris, serve as a site for dystrophic calcification. This deposition of calcium can be readily visualized by radio imaging modalities and hence serves as a prognosticator of the disease process.^1^

Coronary calcium scoring has been recommended by various guidelines for risk stratification, specifically in the setting of primary prevention in asymptomatic individuals. In symptomatic patients, the presence of coronary calcium is associated with future cardiovascular events in the stable chest pain setting and low likelihood of acute coronary syndrome in patients with acute chest pain. However, this information is not routinely quantified as it requires expertise, time, and specialized equipment.^2^

Therefore, risk stratification based on coronary artery calcification serves as an ideal problem set for a deep-learning based solution.

## 2) Data and computation

The data used in the training and testing of these models is sourced from the NLST trial dataset, from the National Cancer Institute under an open license.

17 CT thorax scans were annotated using a random forest classifier and were augmented to 51 volumes (129 in original paper)

The model was built and trained on Google Colab, using an Nvidia V100 GPU. All models were trained for 200 epochs, with batch size 2. Learning rate reduction and early stopping were applied.

## 3) Deep Learning Model

The system consists of three consecutive steps for (1) heart localization, (2) coronary calcium segmentation, and (3) calcium score calculation. Varying configurations of the UNet architecture were trained to perform semantic segmentation in all steps. The respective model architectures used are carried over per se in all steps.

### a) Pre-processing

i. All CT scans were initially padded and cropped to the same size of 512 × 512 × 512pixel (px) and resampled to the same resolution of 0.7 × 0.7 × 2.5 mm/px.

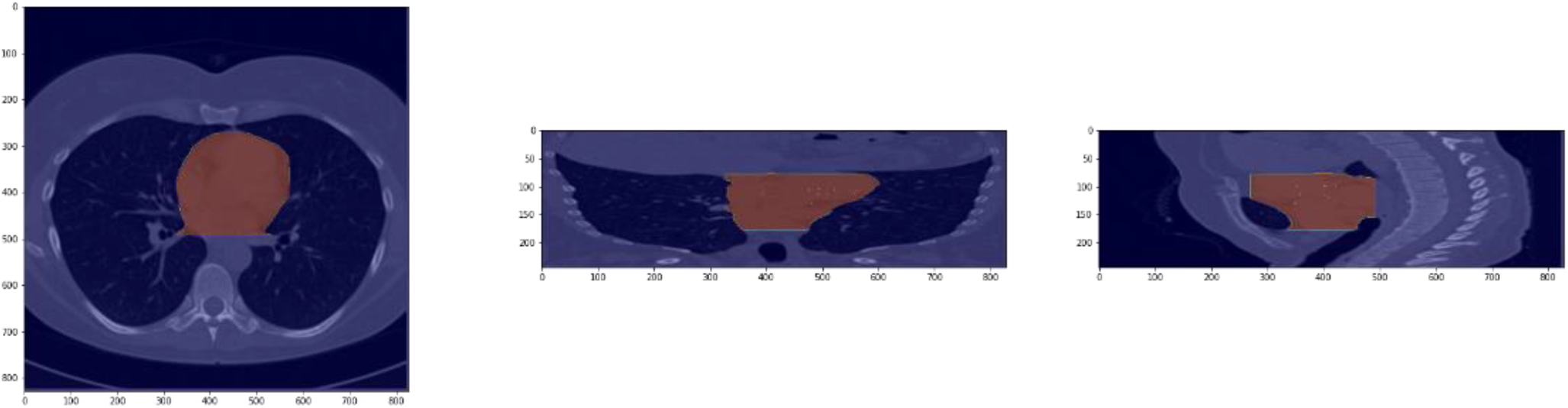

### b) Heart Localization and Segmentation

The first network in our system was trained to localize the heart within a given 3D CT scan. The training cohort was split 80/20 for training and tuning, and all scans were down sampled to a size of 128×128128 to fit into the GPU memory. The model was trained using – UNet with batch normalization, Transfer learning based UNet with Resnet50 and VGG16as encoder and UNet with Pretrained Autoencoder weights for the encoding arm. The autoencoder based model had the highest val-IOU score and f1 score.

The output of the network was up sampled to the initial CT scan size.

### c) Coronary artery segmentation and calcium scoring

For this step, the segmented heart is divided into smaller cubes of size 48 × 48 × 32px. The cubes an overlap to account for adequate representation of low frequency features. The resulting segmentation patches are aligned again, leading to a coronary calcium segmentation of the heart. The final step is to threshold the whole segmentation by 0.95 to obtain the binary calcium mask. The coronary calcium score was derived using a volumetric approach based on the methodology established by Agatston and Janowitz. This involved calculating the score by multiplying the volume of each coronary calcification by a weight factor that corresponds to the highest density observed within the calcification, measured in Hounsfield Units (HU). The weight factors are defined as follows: a factor of 1 for densities between 130 and 199 HU, 2 for densities from 200 to 299 HU, 3 for 300 to 399 HU, and 4 for densities of 400 HU and above. The total calcium score for each patient was obtained by summing the weighted values of all identified calcifications. For further analysis, the calcium risk scores were categorized into four distinct risk groups: very low (0), low (1–100), moderate (101–300), and high (>300).

### d) Data Augmentation

was done by applying Elastic transformation, 3d rotation of 10 degrees, Gaussian blur and Random Gamma.

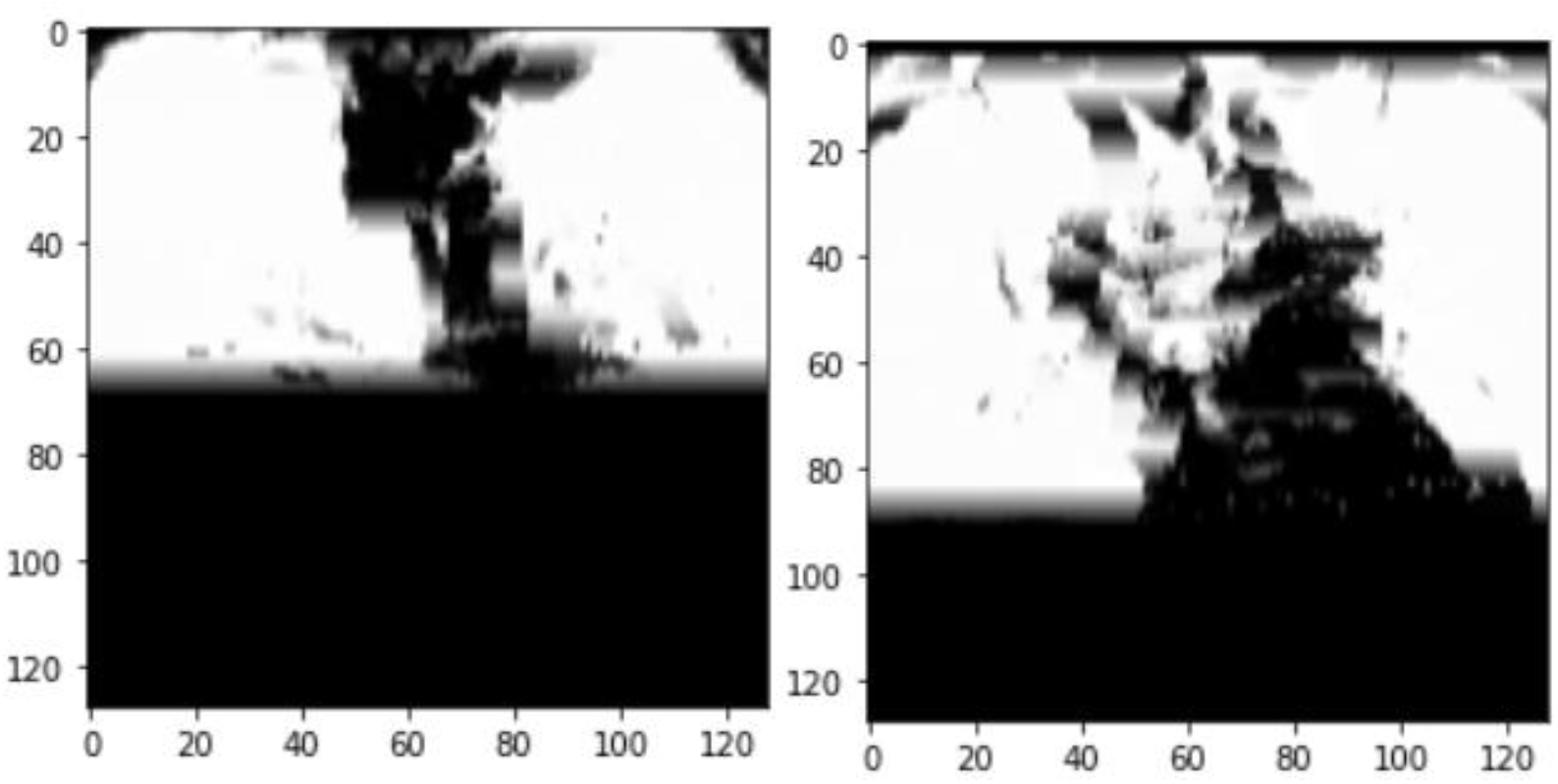

### e) Loss function

used for UNet is a linear combination of Dice loss and Focal loss over image and mask IOU. Equal weights (0.5) were assigned to both components to preserve minority class representation during training.

i. Dice loss given by –

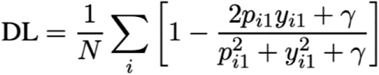
ii. Focal loss given by –

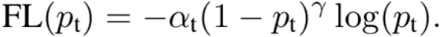

### f) Activation function used is Scalar Exponential Linear Unit(SELU)

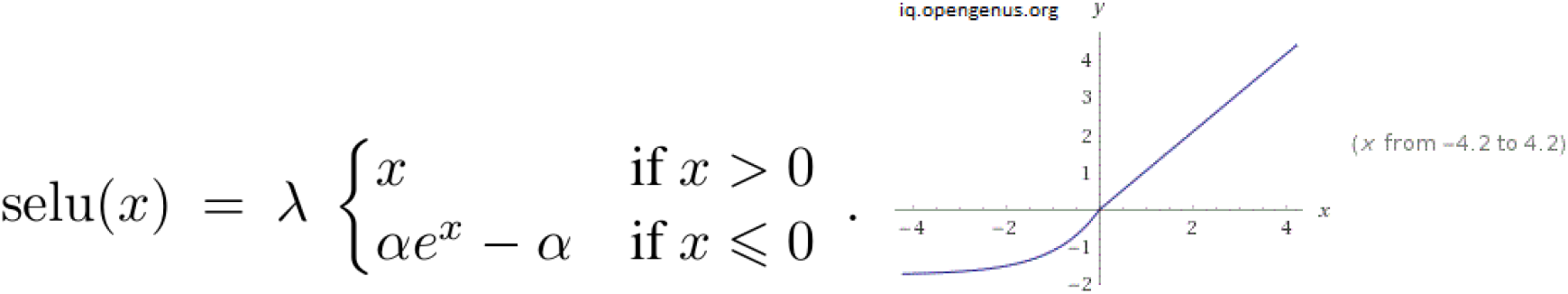

i. SELU was employed to fix the ‘dying ReLU” problem from negative weights/biases forcing the derivative to go to 0 and causing arrest of weight updation and neuron activation.
ii. SELU is also self-normalizing, thereby promoting faster convergence of the model towards global minima

### g) Optimizer used is ADAM (Adaptative Learning Rate Optimization Algorithm) for fast convergence

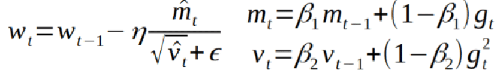

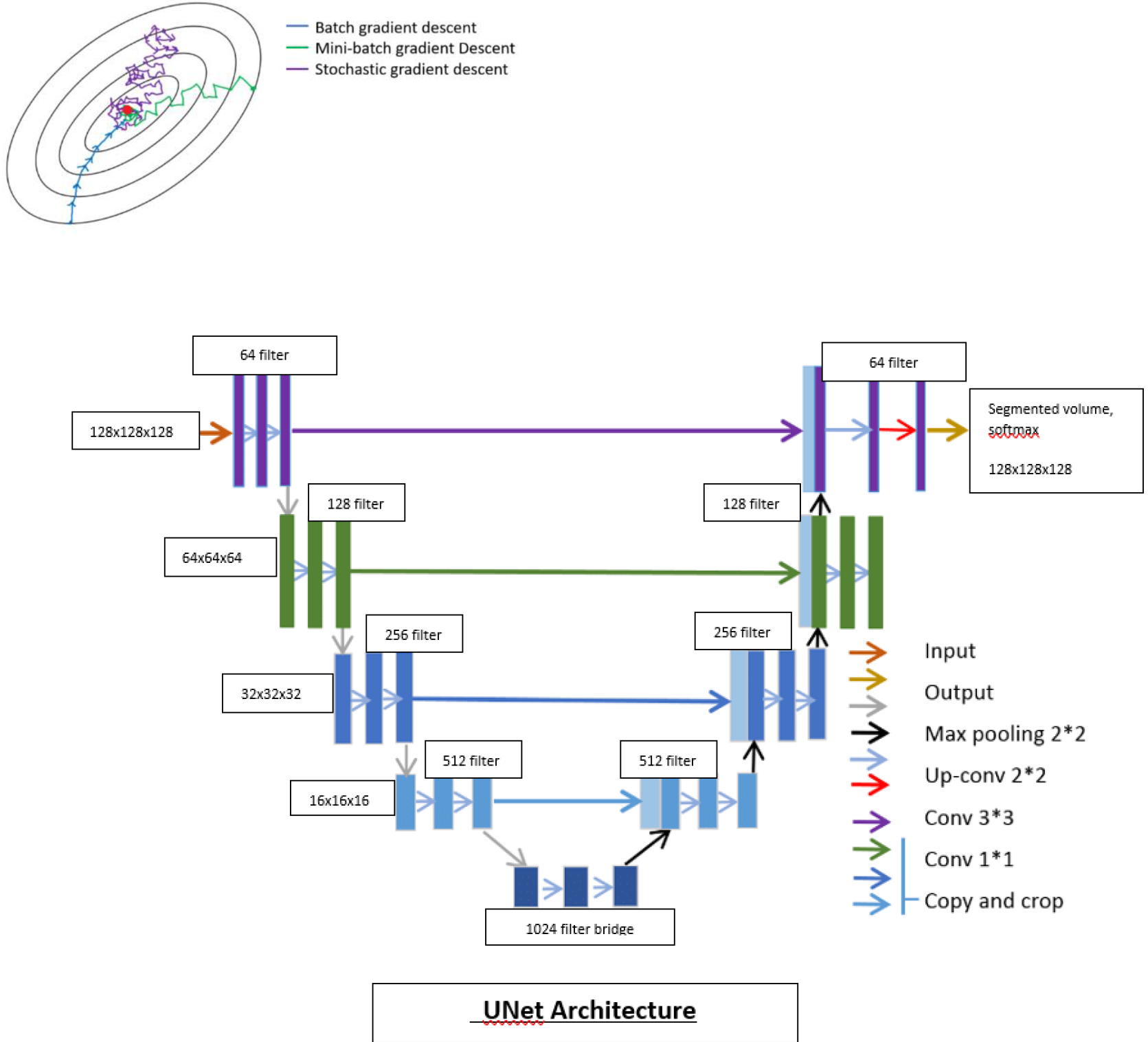

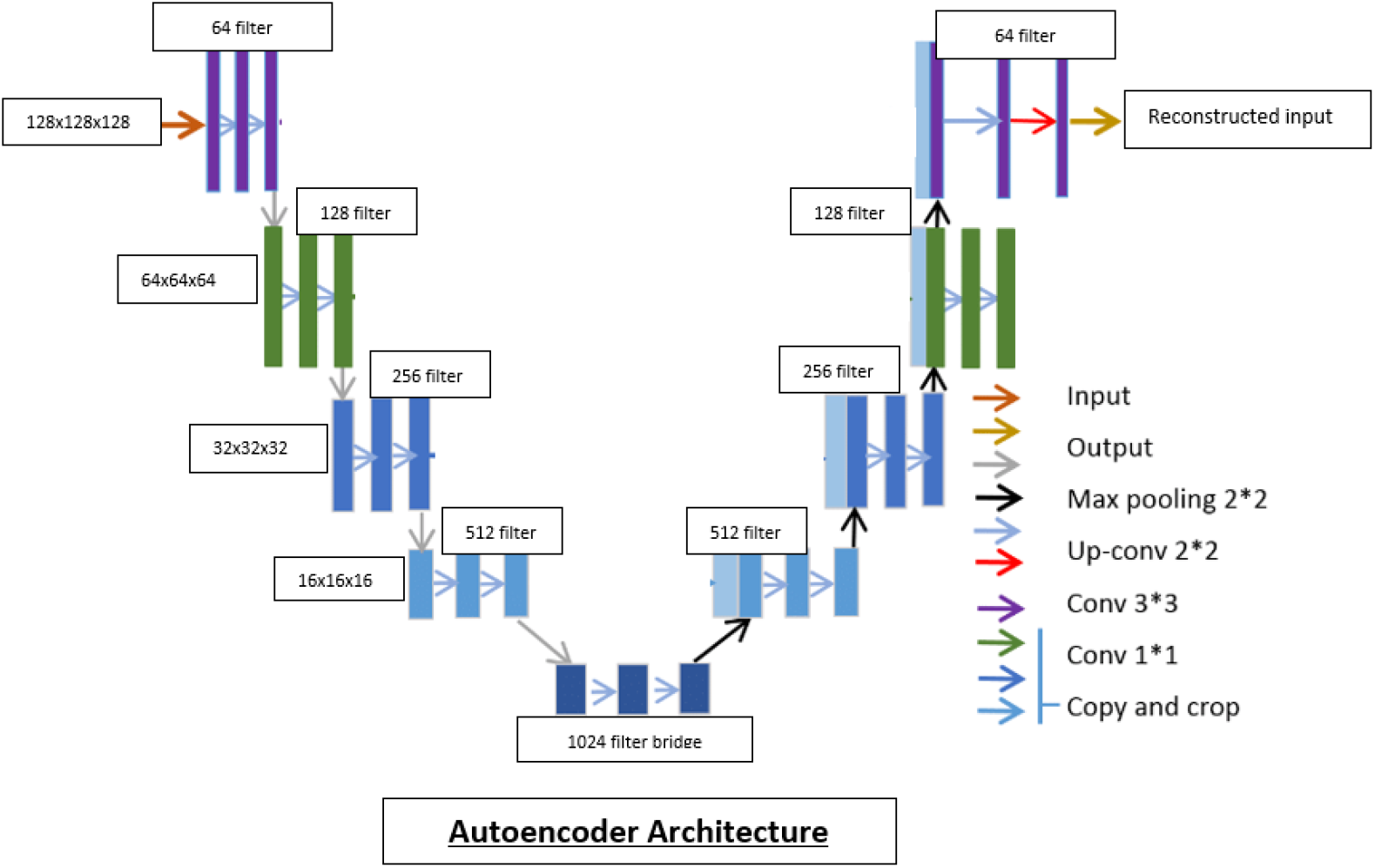

## 4) Results

- The models were evaluated on Dice-Coefficient/IOU (intersection over union) score and f1 score based on the manually segmented masks vs. predicted masks, Dice coefficient given by –

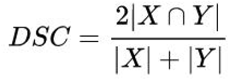

And f1 score by –

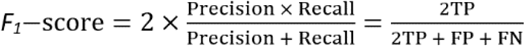
- The IOU’s and f1 scores evaluated for the models are as follows –
  ∘ UNet with batch normalization – 0.53 IOU, 0.48 f1
  ∘ UNet with ResNet 50 encoder – 0.76 IOU, 0.79 f1
  ∘ Extended UNet with Autoencoder – 0.78 IOU, 0.83 f1
- The reference paper quotes a validation IOU score of 0.77 using UNet trained on 189 manually annotated volumes for 1200 epochs, while the autoencoded extended UNet with data augmentation, batch normalization and other optimizations achieved a validation IOU of 0.78 with 21 manually annotated samples and 100 epochs of training.

### a) UNet with batch normalization

- The UNet with batch normalization shows poor learning over 250 epochs, with training data overfitting by the first 50 epochs, with only a rise of 0.1 IOU during training.
- Validation IOU remains poor with under 0.5, clearly indicating overfitting and poor robustness of the model
- 250 epochs were completed in 2hrs 15min of training

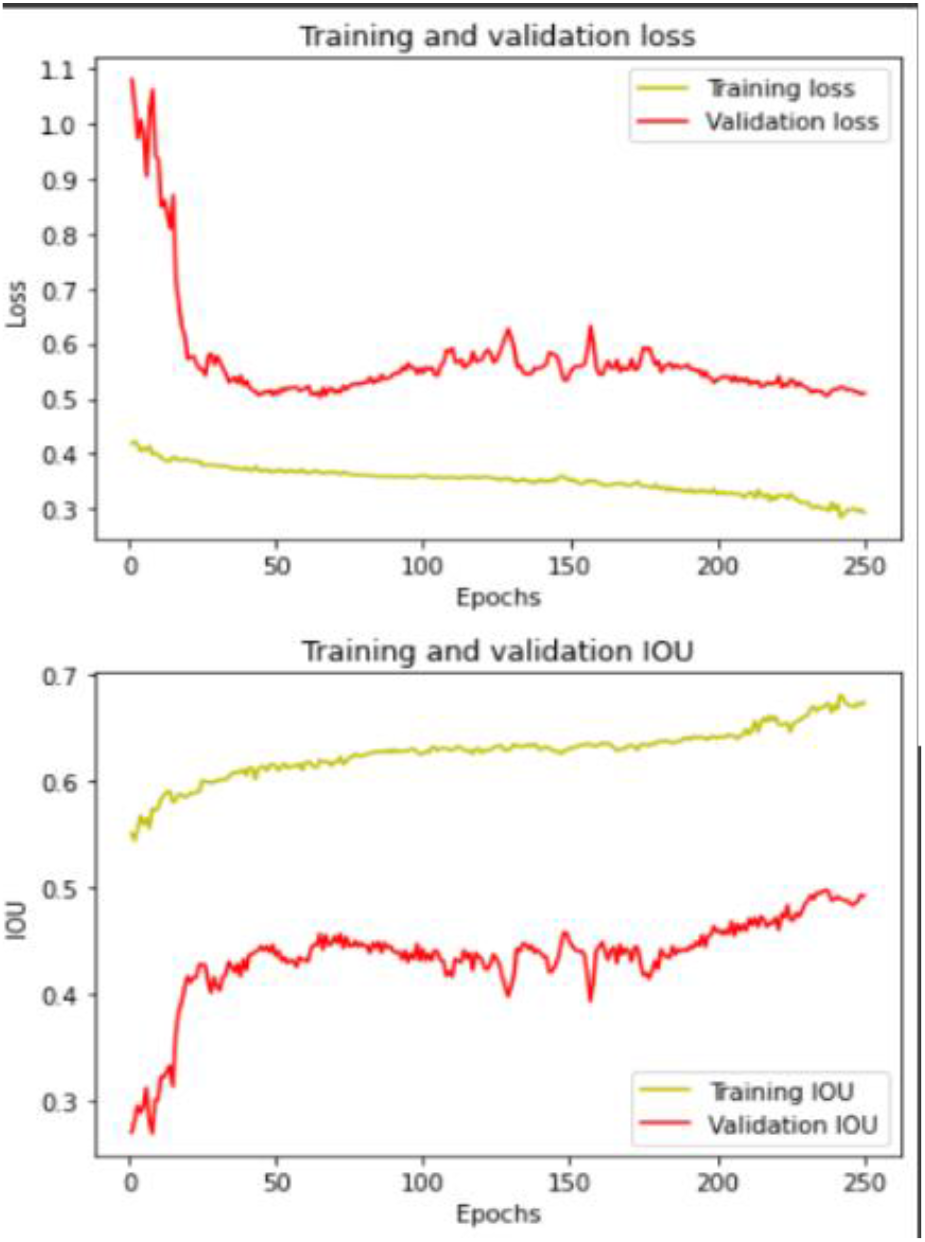

### b) UNet with Autoencoder

- UNet model initialized with pretrained autoencoder weights performed the best, with a validation IOU of 0.78 over 100 epochs of training.
- As expected of an autoencoder backbone trained on similar CT scans, the model was quick to learn and demonstrated a high IOU score withing the first 20 epochs and the low fluctuations in loss over the later epochs show robustness and good generalization
- 100 epochs of training were completed in 50 minutes.

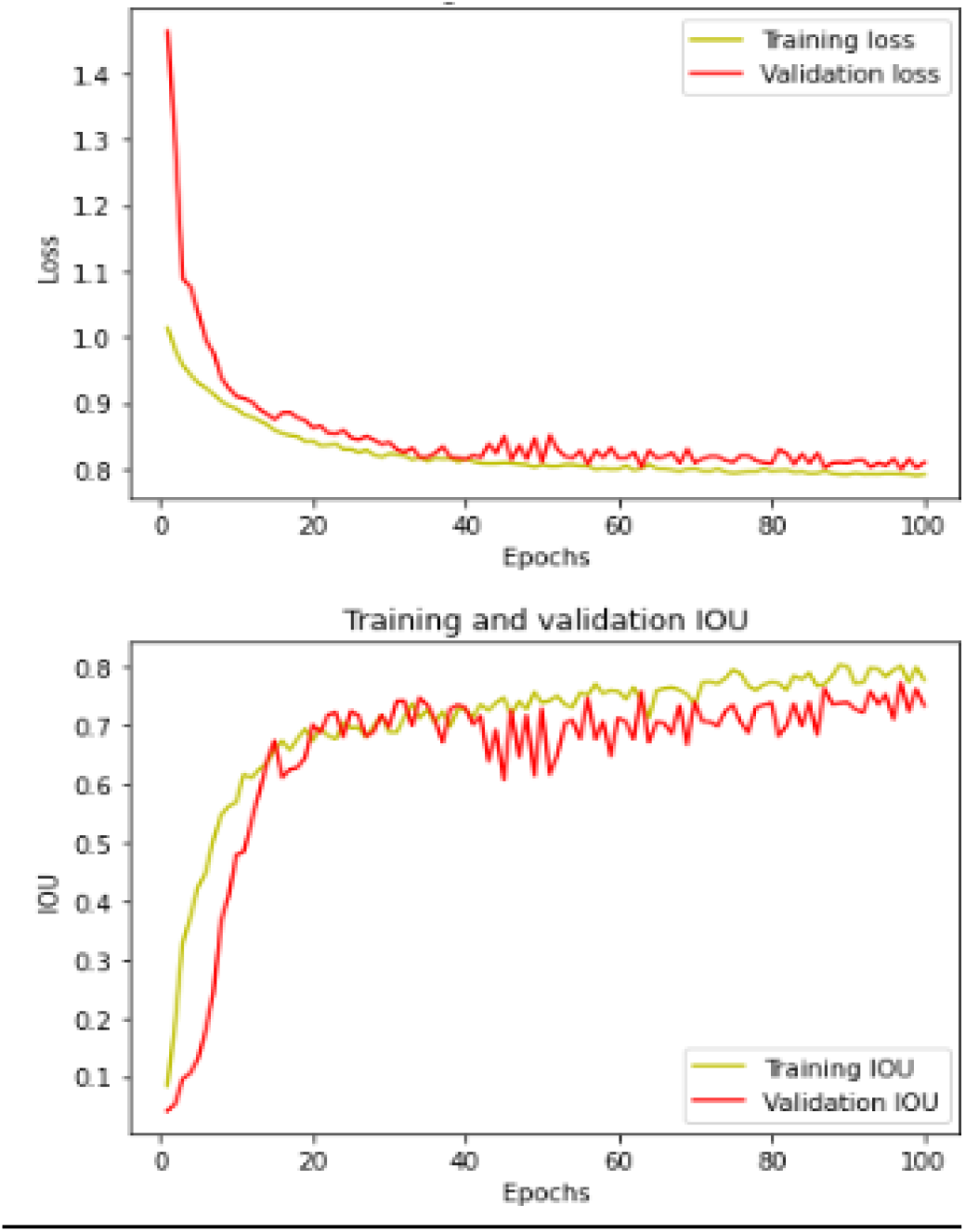

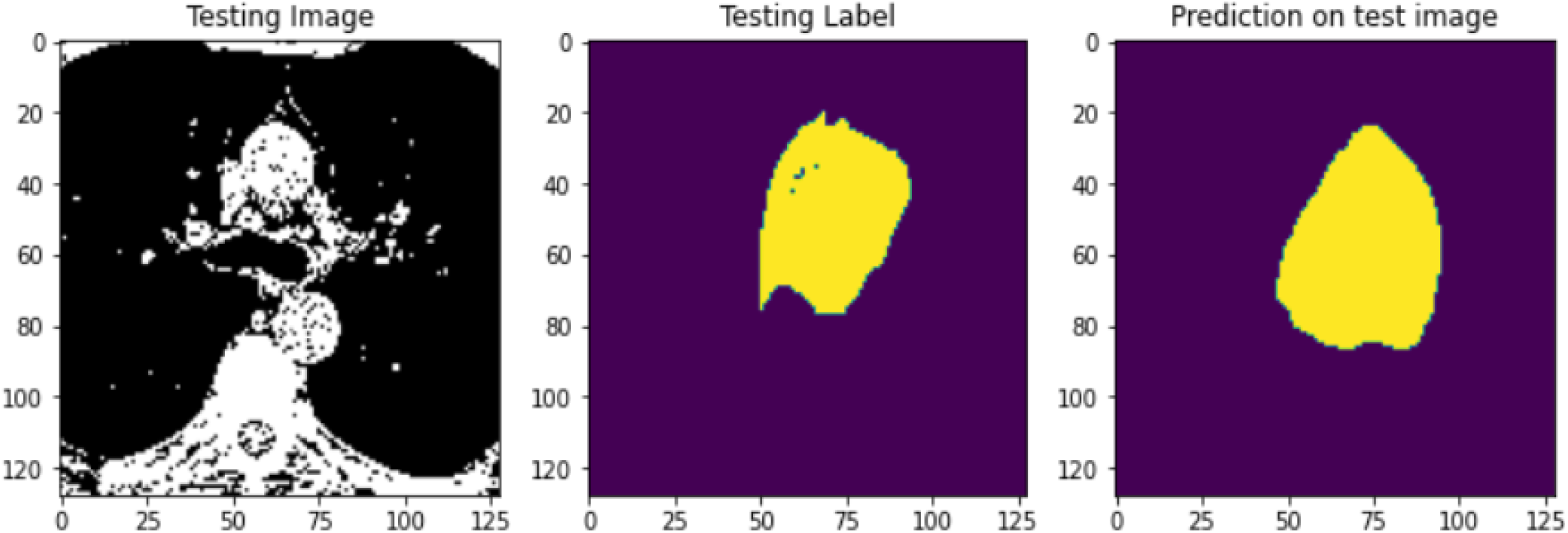

### c) UNet with Resnet-50 encoder arm

- UNet model was trained with multiple encoder backbones – vgg16, renset50, densenet21 and effecientnetb5m with imagenet weights.
- The resnet50 performed the best with 0.76 validation IOU over 80 epochs. Although the model seemed to overfit the dataset, with training IOU approaching 0.92
- Validation IOU also shows high fluctuation during training
- 80 epochs of training completed in 55 minutes.

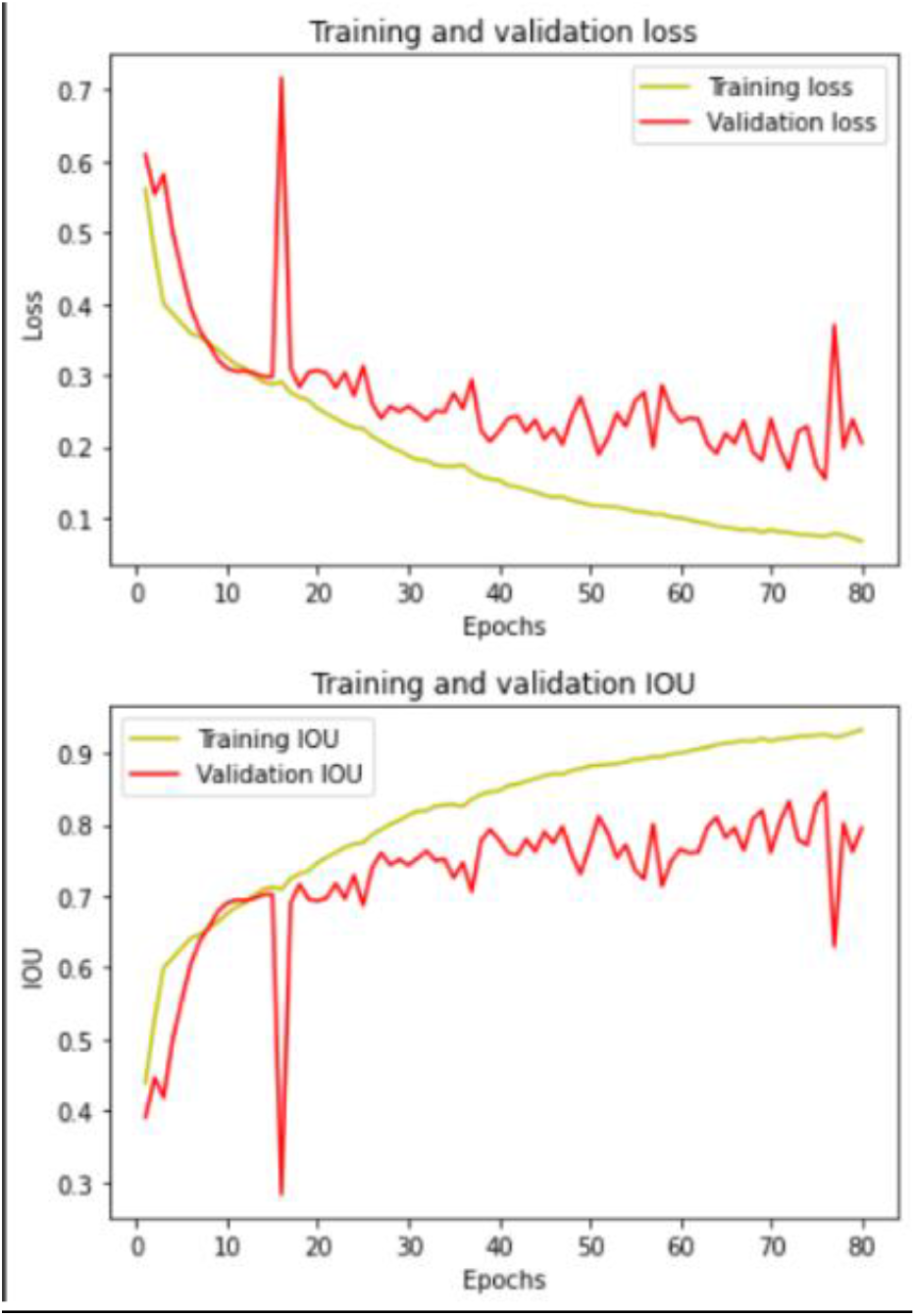

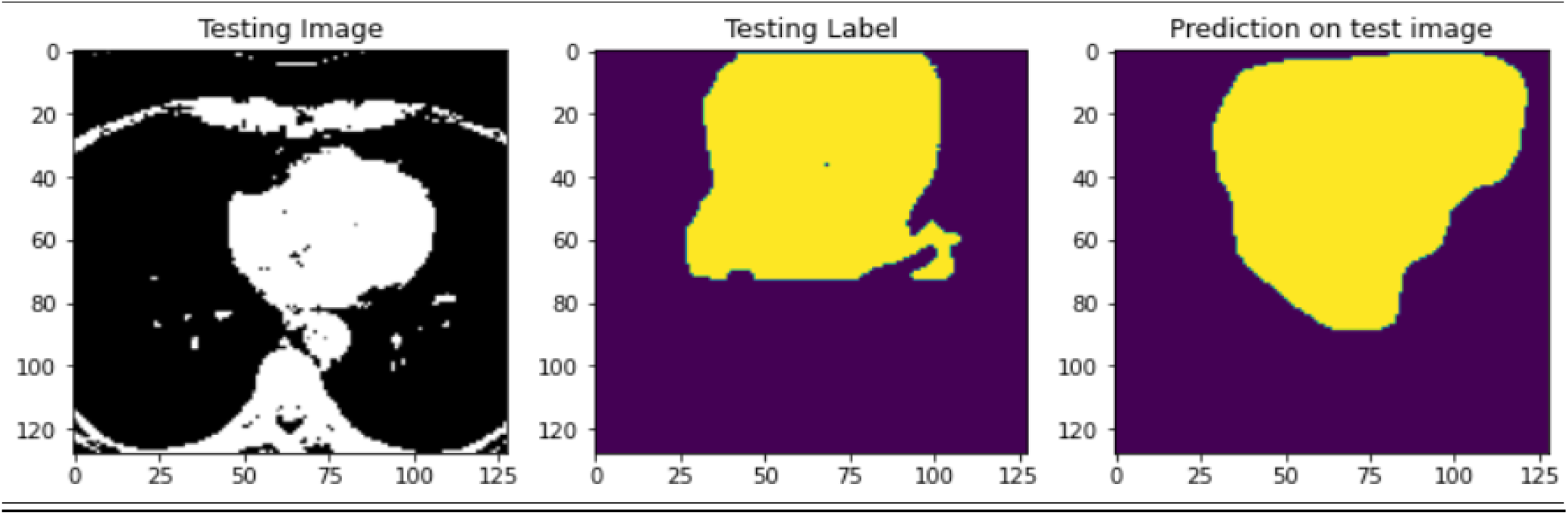

## Discussion

In this project, I have tried to demonstrate a computationally efficient and robust deep learning algorithm to segment and predict the severity and incurred risk of coronary artery disease using a less-than-ideal dataset, with fewer CT volumes and masks. Mask and risk prediction was robust with respect to ground truth even in volumes with poor scanning/annotation quality. People with a calcium score of zero are at very low risk, and risk increases as the calcium score increases, according to the deep learning system’s determined ordinal calcium score tiers. According to the 2018 ACC/AHA^3^ guidelines, a calcium score of 0 indicates very low risk and unlikely benefit from statin therapy, whereas a high calcium score (100 or 75th centile for age/sex) indicates that a statin should be taken into consideration in people at intermediate risk (defined as 7.5 percent to 19.9 percent 10-year risk of cardiovascular events based on risk factors). Despite these recommendations, a dedicated coronary calcium scoring CT is not yet covered by Medicare and most US insurance companies, and for this reason, there is a great deal of interest in deriving the calcium score from routine chest CTs, which are far more common.

Traditionally, coronary artery segmentation and 3d calcium scoring requires clinical and technical expertise and the use of special software capable of handling such tasks, making It highly inefficient and impractical as a screening tool for heart disease. The deep learning based automated calcium scoring method addresses this concern by reliably and accurately extracting the calcium score in both cardiac CT and chest CT. The system calculates the calcium score in under 2 s, without human input.

Although other studies have investigated deep learning algorithms for automated coronary calcium quantification, they used inefficient computation with models showing poor robustness when compared with the size of the dataset/complexity of training. For example, previous publications for fully automatic coronary calcium assessment used CNNs for absolute risk assessment without semantic segmentation, leading to poor interpretability^4,5^. Shadmi et al.^6^ proposed a 2d FC-Densent and UNet architecture that performed well on a small section of NLST trial data. The study inspiring this method by Roman et al^7^. employs successive 3D Unet models to sequentially segment the heart, coronary arteries and extract calcium scoring values. However, the model shows poor performance when trained on smaller datasets with improper annotations, leading to poor generalizability and low accuracy. This method iterates on the original by making strides in data augmentation, computational efficiency and robustness on imperfect datasets by employing more modern advancements like-ML accelerated annotation of training data, autoencoders, transfer learning and data augmentation. This method shows comparable accuracy to the original while only using a fraction of the resources and training time.

Further improvements to this study could be made in the aspects of-

- Data augmentation – Employing Generative Adversarial Networks to generate Scan-Mask pairs
- More robust architectures by using Residual blocks and Attention layers to maximize both spatial and pixel awareness
- Explore other methods of automated annotation of training data

In summary, the end-to-end deep learning system provides an automated quantification of coronary calcium on both cardiac CT and lung cancer screening CT. The deep learning calcium score is strongly associated with cardiovascular risk in a broad spectrum of clinical scenarios. Automated quantification of coronary calcium has the potential to improve clinical routine and population health.

## Data Availability

All data produced in the present study are available upon reasonable request to the authors

## Author approval

All authors have reviewed and approved the final version of this manuscript. The sole author confirms that there are no conflicts of interest related to this study and is committed to maintaining the integrity of the research process.

## Code/Data availability

https://github.com/MedMachine00/Deep-learning-based-coronary-artery-segmentation

